# Accelerated brain age in young to early middle-aged adults after mild to moderate COVID-19 infection

**DOI:** 10.1101/2024.03.05.24303816

**Authors:** Shelli R Kesler, Oscar Y. Franco-Rocha, Alexa De La Torre Schutz, Kimberly A. Lewis, Rija M Aziz, W. Michael Brode, Esther Melamed

## Abstract

Cognitive decline is a common adverse effect of the Coronavirus Disease of 2019 (COVID-19), particularly in the post-acute disease phase. The mechanisms of cognitive impairment after COVID-19 (COGVID) remain unclear, but neuroimaging studies provide evidence of brain changes, many that are associated with aging. Therefore, we calculated Brain Age Gap (BAG), which is the difference between brain age and chronological age, in a cohort of 25 mild to moderate COVID-19 survivors (did not experience breathlessness, pneumonia, or respiratory/organ failure) and 24 non-infected controls (mean age = 30 +/− 8) using magnetic resonance imaging (MRI). BAG was significantly higher in the COVID-19 group (F = 4.22, p = 0.046) by 2.65 years. Additionally, 80% of the COVID-19 group demonstrated an accelerated BAG compared to 13% in the control group (X^2^ = 20.0, p < 0.001). Accelerated BAG was significantly correlated with lower cognitive function (p < 0.041). Females in the COVID-19 group demonstrated a 99% decreased risk of accelerated BAG compared to males (OR = 0.015, 95% CI: 0.001 to 0.300). There was also a small (1.4%) but significant decrease in risk for accelerated BAG associated with longer time since COVID-19 diagnosis (OR = 0.986, 95% CI: 0.977 to 0.995). Our findings provide a novel biomarker of COGVID and point to accelerated brain aging as a potential mechanism of this adverse effect. Our results also offer further insight regarding gender-related disparities in cognitive morbidity associated with COVID-19.

## Introduction

Emerging studies have demonstrated that the Coronavirus Disease of 2019 (COVID-19) increase the risk for a range of neurological difficulties including cognitive impairment. Cognitive decline, or “brain fog”, is believed to be part of a cluster of symptoms that encompass difficulty with concentration, attention, and memory that persist beyond the acute disease phase. These symptoms are known as Post-acute Sequelae of SARS-CoV-2 Infection (PASC) or Long COVID (1). Cognitive impairment after COVID-19 (COGVID) reduces quality of life, extends disease related morbidity, and impairs ability to return to work (2, 3). The precise mechanism of COGVID is unclear though neuroimaging studies have demonstrated significant alterations in brain structure and function (4–13), including studies of non-critical/non-hospitalized cases (14–17).

We previously showed that survivors of mild to moderate, non-hospitalized COVID-19 have significantly lower functional brain connectivity compared to non-infected controls, and hypo-connectivity was correlated with lower cognitive performance (18). Prior studies have indicated that reduction in functional brain connectivity is associated with normal aging (19, 20). Given that our cohort of COVID-19 survivors was young (mean age = 30 years), accelerated brain aging may be a potential mechanism of cognitive impairment in this population. Prior studies of COGVID have focused on older adults with severe disease as these are the most vulnerable patients. However, this approach has created a gap in the literature regarding cognitive outcomes in survivors of mild to moderate COVID-19 (people that do not experience breathlessness, pneumonia, or respiratory/organ failure), as younger individuals also show significant COGVID and may in fact be more vulnerable to long-term impairment (18, 21, 22).

Brain age can be estimated using non-invasive neuroimaging in combination with artificial intelligence. Comparing estimated brain age with chronological age provides a metric known as Brain Age Gap (BAG), which quantifies brain health as the divergence from the typical trajectory of aging (23, 24). Multiple studies have shown that BAG is a sensitive biomarker for detecting various neurologic and neuropsychiatric conditions (25, 26). We aimed to examine BAG in young and early middle-aged adults who had mild to moderate COVID-19. We hypothesized that BAG would be higher in COVID-19 survivors compared to non-infected controls.

## Methods

### Participants

Between October 2021 and January 2023, we recruited adults with and without history of COVID-19 in central Texas. Potential participants were recruited via social media, community boards, and outpatient referrals. Participants in the COVID-19 group were excluded for signs or symptoms of severe infection including self-rating of severity or hospitalization. Any potential participant was excluded for pre-existing history of developmental, medical, or psychiatric disorders known to affect cognitive function, significant sensory impairment (e.g., blindness), or MRI contraindications (e.g., magnetic biomedical implants, certain orthodontia, claustrophobia). In total, we enrolled 50 adults (54% female) aged 21 to 50 years (mean = 30.7 +/− 8.7). Twenty-six participants had a self-reported history of positive COVID-19 test and the remaining 24 participants self-reported no history of infection by test or associated symptoms. We excluded one participant in the COVID-19 group for receiving treatment in a Post-COVID Clinic for Long COVID. There were no significant differences between the two groups in any demographic characteristics except for racial/ethnic minority status which was significantly higher in the control group (Table 1). This study was approved by the University of Texas at Austin Institutional Review Board (protocol# 00001337), was conducted in accordance with the Declaration of Helsinki, and all participants provided written, informed consent.

**Table 1.** Demographic and COVID-19 Characteristics. Data are shown as mean (standard deviation) unless otherwise indicated.

|  | **COVID N=25** | **Control N=24** | **stat** | **p value** |
| --- | --- | --- | --- | --- |
| Age (years) | 30.3 (8.0) | 30.3 (9.0) | t = -0.003 | 0.997 |
| Education (years) | 16.4 (1.8) | 16.9 (1.6) | t = 0.995 | 0.325 |
| Female gender | N = 14 (56.0%) | N = 13 (54.2%) | $\chi^2 = 0.017$ | 0.897 |
| Income < \$100K | N = 12 (48%) | N = 8 (33.3%) | $\chi^2 = 1.113$ | 0.292 |
| Racial/ethnic minority | N = 3 (12.0%) | N = 11 (45.8%) | $\chi^2 = 6.868$ | 0.009 |
| Time since diagnosis (days) | 307 (223)<br>range: 12-719 |  |  |  |
| COVID severity moderate | N = 11 (44.0%) |  |  |  |
| No. of symptoms during active COVID infection | median = 8.0,<br>IQR = 4.6<br>range = 1-16 out of 20 possible |  |  |  |

### Demographic and COVID-19 Measures

We used instruments recommended by the National Institutes of Health to facilitate COVID-19-related research. Specifically, from the NIH Repository of COVID-19 Research Tools, we administered the RADxUP Sociodemographic Questionnaire and the COVID-19 Experiences questionnaire to measure COVID-19 severity and symptoms during active infection (27). We also assessed current anosmia using the Pocket Smell Test (28).

### Neuropsychiatric Function

We administered BrainCheck, a computerized battery of neuropsychological tests including Trails A (attention, processing speed, executive function), Trails B (attention, processing speed, executive function), Immediate List Recall (verbal memory learning), Delayed List Recall (verbal memory recall), Stroop (response inhibition), and Digit Symbol (graphomotor processing speed) (29). BrainCheck provides age normalized test scores (mean = 100 +/− 15). To measure subjective cognitive function, we administered the Patient Reported Outcome Measures Information System (PROMIS) Cognitive Function Short Form 8a (30). This is an 8-item, self-rating questionnaire regarding the frequency of cognitive symptoms. We also administered PROMIS-57 Profile v2.1 to measure depressive symptoms, fatigue, anxiety, sleep disturbance, pain, and social role functioning (31). PROMIS provides standardized T-scores with a mean of 50 and standard deviation of 10.

### Neuroimaging Pulse Sequence

T1-weighted anatomical MRI was collected using a high-resolution magnetization prepared rapid gradient echo (MPRAGE) sequence: TR = 2400ms, TE = 2.18, flip angle = 8 degrees, FOV 256 mm, parallel imaging with GRAPPA acceleration factor = 2, 0.8mm isotropic resolution, 208 sagittal slices, scan time = 6:38min. Resting state fMRI, diffusion tensor imaging and arterial spin labeling data were also collected during this 45-minute scan session but are not reported here.

### Brain Age Gap (BAG)

We estimated brain age from brain volumes with brainageR v2.1, a publicly available algorithm that has been shown to be one of the most reliable for predicting age from brain MRI (32). The algorithm implements a Gaussian Processes regression model to predict brain age from segmented brain volumes (33, 34). The model was trained on 3,377 healthy individuals (mean age = 40.6 years, SD = 21.4, age range 18-92 years) and tested on an independent dataset of 857 healthy individuals (mean age = 40.1 years, SD = 21.8, age range 18-90 years). The algorithm accepts raw, T1-weighted MRI scans, segments and normalizes them using Statistical Parametric Mapping v12 (Wellcome Trust Centre for Neuroimaging, London, UK) with custom templates. We subtracted chronological age from estimated brain age to calculate BAG. A positive BAG thus represents accelerated brain age and a negative BAG represents decelerated brain age (23).

### Statistical Analysis

We compared BAG between groups using two different approaches; ANCOVA controlling for total brain volume, and a Chi square test to evaluate the difference in frequency of accelerated BAG (BAG > 0). We conducted exploratory Spearman correlation analysis between accelerated BAG (1 = yes, 0 = no) and cognitive test scores. We used exploratory logistic regression to determine clinical and demographic characteristics (number of days since COVID-19 diagnosis, number of COVID-19 symptoms, sex, COVID-19 severity, and racial/ethnic minority classification) associated with accelerated BAG. Cognitive tests were compared using t-tests for continuous variables and Chi Squared tests for categorical variables during our previously study of this cohort with false discovery rate (FDR) correction for multiple comparisons (18). Alpha level for all tests was set at p < 0.05 and all data visualizations and analyses were conducted in the R Statistical Package v4.3.1 (R Foundation, Vienna, Austria).

## Results

### COVID-19 Group Characteristics

As we previously reported for this cohort, participants with a history of COVID-19 infection were nearly evenly split between mild and moderate disease severity with relatively low symptom burden during the acute infection (18). There was a wide range in the interval between COVID-19 diagnosis and study enrollment (12-719 days, mean of 10 +/− 7 months, Table 1). No participants demonstrated anosmia (Table 2).

**Table 2.** Neuropsychiatric Testing Performance. Data are shown as mean (standard deviation). Rank biserial correlation is the effect size. FDR = false discovery rate correction for multiple comparisons.

|  | **COVID**<br>**N=25** | **Control N**<br>**= 24** | **stat**<br>**(W/X<sup>2</sup>)** | **uncorrected**<br>**p value** | **FDR**<br>**corrected**<br>**p value** | **rank biserial**<br>**correlation** |
| --- | --- | --- | --- | --- | --- | --- |
| Trails A | 97.2<br>(11.9) | 104.4<br>(13.7) | 372.5 | 0.081 | 0.211 | 0.296 |
| Trails B | 98.6<br>(11.9) | 100.8<br>(15.5) | 333.0 | 0.353 | 0.386 | 0.158 |
| Digit symbol<br>substitution | 95.0<br>(14.8) | 99.2 (15.1) | 353.5 | 0.289 | 0.386 | 0.178 |
| Stroop | 94.4<br>(19.4) | 101.1<br>(15.8) | 341.5 | 0.269 | 0.386 | 0.188 |
| Immediate<br>recall | 100.2<br>(18.9) | 106.5 (9.8) | 344.0 | 0.356 | 0.386 | 0.147 |
| Delayed recall | 102.4<br>(13.8) | 106.5<br>(10.5) | 359.5 | 0.219 | 0.386 | 0.198 |
| PROMIS<br>cognitive | 49.4<br>(11.4) | 60.1 (6.2) | 475.0 | 0.000 | < 0.001 | 0.583 |
| Impaired<br>PROMIS<br>cognitive (T<br>score < 55) | 18 (72%) | 6 (25%) | 10.8 | 0.001 | 0.007 | - |
| PROMIS social<br>role<br>performance | 57.2 (9.7) | 59.1 (8.4) | 332.0 | 0.494 | 0.494 | 0.107 |
| PROMIS<br>anxiety | 55.0<br>(11.6) | 48.0 (9.4) | 195.0 | 0.035 | 0.199 | -0.350 |
| PROMIS<br>depression | 48.9 (8.9) | 43.7 (5.4) | 202.5 | 0.046 | 0.199 | -0.325 |
| PROMIS<br>fatigue | 49.9 (9.9) | 44.4 (9.2) | 212.0 | 0.078 | 0.211 | -0.293 |
| PROMIS sleep<br>disturbance | 50.9 (9.8) | 46.5 (7.2) | 236.0 | 0.202 | 0.386 | -0.213 |
| PROMIS pain | 45.5 (7.1) | 43.8 (6.0) | 260.0 | 0.333 | 0.386 | -0.133 |
| Anosmia<br>(Pocket Smell<br>Test score < 2) | 0 (0%) | 0 (0%) | - | - | - | - |

### Neuropsychiatric Function

The COVID-19 group also demonstrated lower performance on all cognitive tests (effect size r = 0.15 to 0.30) (18). However, only PROMIS Cognitive score was significantly different between groups (p < 0.001, FDR corrected, effect size r = 0.58) and the COVID-19 group demonstrated significantly higher incidence of PROMIS Cognitive impairment (X^2^ = 10.8, p = 0.007, FDR corrected). The COVID-19 group also endorsed greater symptoms of anxiety and depression; however, these were not significantly different between groups after correction for multiple comparisons (Table 2).

### Between Group Difference in BAG

As shown in Figure 1, mean BAG was significantly higher in the COVID-19 group (F = 4.22, p = 0.046) by 2.65 years. Additionally, there were N = 20 (80%) in the COVID-19 group who demonstrated an accelerated BAG compared to N = 3 (13%) in the control group, which was a significant difference (X^2^ = 20.0, p < 0.001).

**Figure 1.**
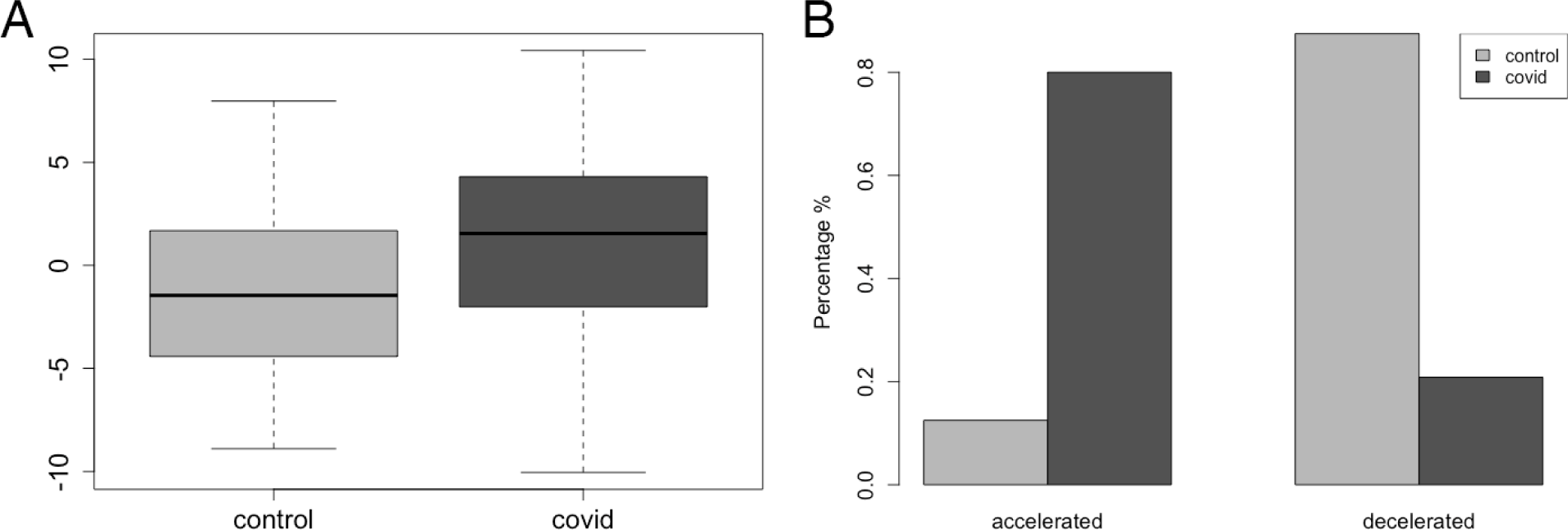
Brain Age Gap (BAG) A: BAG was significantly higher in the COVID-19 group compared to non-infected controls (p = 0.046). B: Accordingly, the proportion of individuals with accelerated BAG (BAG > 0) was significantly higher in the COVID-19 group (p < 0.001).

### Relationship Between Accelerated BAG and Cognitive Function

Accelerated BAG in the COVID-19 group was associated with lower Trails A performance (r = - 0.348, p = 0.041) as well as PROMIS Cognitive impairment (r = −0.306, p = 0.033), but no other cognitive test scores (p > 0.127).

### Clinical and Demographic Characteristics of Accelerated BAG

The overall logistic regression model was significant (X^2^ = 14.71, Nagelkerke R^2^ = 0.666, p = 0.012, Table 3). An increased duration from COVID-19 diagnosis was associated with a small yet statistically significant decrease (1.4%) in the risk of accelerated BAG. There was also a large (99%) and significant decreased risk of accelerated BAG in females compared to males with COVID-19. There were N = 9/14 females (64%) in the COVID-19 group with accelerated BAG compared to 10/11 males (91%). Increased number of symptoms during acute infection, moderate COVID-19 severity, and racial/ethnic minority status were associated with increased risk for accelerated BAG but were not statistically significant and lacked precision in this small sample (i.e., large confidence intervals for odds ratios).

**Table 3.** Clinical and Demographic Characteristics of Accelerated Brain Age Gap (BAG) The logistic regression model was significant (X^2^ = 14.71, Nagelkerke R^2^ = 0.666, p = 0.012).

|  | **Odds Ratio** | **95% CI lower** | **95% CI upper** |
| --- | --- | --- | --- |
| Time since COVID diagnosis (days) | 0.986 | 0.977 | 0.995 |
| Number of COVID symptoms* | 1.35 | 0.873 | 2.09 |
| Female biological sex | 0.015 | 0.001 | 0.300 |
| Moderate COVID severity | 6.82 | 0.088 | 526 |
| Racial/ethnic minority | 31.52 | 1.55 | 642 |

\*During acute infection

## Discussion

Cognitive impairment after COVID-19 (COGVID) appears to be one the most common long-term side effects of infection (1) yet the mechanisms of this problem are an area of active research. In this study, we identified accelerated brain aging as a potential contributor to COGVID. Individuals with a history of COVID-19 infection demonstrated a 2.65-year accelerated BAG compared to the control group. We also found that poorer performance on an objective measure of attention, processing speed, and executive function (Trails A), as well as greater subjective cognitive problems were associated with higher BAG. Finally, we identified a significantly higher risk of accelerated BAG among males compared to females with mild to moderate COVID-19 infection.

COVID-19 induced upregulation of angiotensin-converting enzyme 2 (ACE2) expression in the brain is a potential mechanism for our finding of accelerated BAG. SARS-CoV-2, the virus responsible for COVID-19, uses ACE2 receptors to enter human cells (35). Furthermore, ACE2 expression is upregulated in the brain after COVID-19, especially in patients with significant neurologic symptoms (36). ACE2 is primarily involved in the renin-angiotensin system (RAS). While the RAS is primarily known for its role in regulating blood pressure and fluid balance, it is also crucial for brain health and cognitive function (37). The RAS system becomes dysregulated with increasing age, contributing to the pathogenesis of neurodegenerative diseases (38, 39). The specific role of ACE2 in neurodegeneration is complex given that its expression is associated with neuroprotective effects, yet several studies have noted elevated ACE2 levels in patients with Alzheimer’s disease (40–42). These studies suggest that chronic elevation of ACE2 may result in dysregulation of the RAS system, increasing the risk for accelerated brain aging. To further evaluate this hypothesis, future studies should assess ACE2 level in Long COVID patients with cognitive impairment.

Alternatively, the impact of ACE2 expression may vary depending on its distribution across different brain regions (43). Some areas might be more sensitive to changes in ACE2 levels, leading to localized neurodegeneration despite the overall neuroprotective effects of ACE2. Our group and others have shown that prefrontal cortex is preferentially susceptible to COVID-19 (9, 14, 15, 18). Under normal conditions, ACE2 expression is extremely low in prefrontal cortex (44) and thus this region may be exceptionally vulnerable to ACE2 elevation. Currently available brain age algorithms do not provide regional BAGs, but these could be estimated using custom models in large samples.

COGVID may also result from accelerated aging due in part to chronic inflammation, or “inflammaging” (45, 46). Inflammation is well-known to play a critical role in neurodegeneration (47, 48). Patients with COVID-19 demonstrate post-infection upregulation of inflammatory biomarkers (49, 50) with the extent of the inflammatory response depending on disease severity (49, 50). Research among middle-aged COVID-19 survivors has shown significant inflammation-related astrocytic damage and neural dysfunction regardless of disease severity (51). Inflammation after COVID-19 infection may also impair cognition via reduced serotonin levels (52). The prefrontal cortex is particularly vulnerable to inflammation due to its unique reliance on glutamate receptor/calcium mediated neurotransmission (53). The relationship between inflammation and BAG over time in mild and moderate COVID-19 survivors requires further study.

BAG is associated with certain genetic as well as lifestyle factors and may therefore be useful in treatment monitoring and development (23, 54, 55). Dietary and nutritional interventions have been effective in reducing brain aging, particularly calorie restriction and low consumption of processed food and sweets (56, 57). Physical activity has consistently been associated with improved cognitive function and decreased brain aging (58, 59). However, using physical activity to treat Long COVID is currently highly controversial in clinical practice because of the risk for inducing post-exertional malaise. There appears to be a complex interaction between exercise and immune-mediated symptoms, especially among younger COVID-19 survivors (49) and therefore, physical activity may not be the best recommendation for many patients. Current guidance on Long COVID emphasizes that physical activity should be individualized and structured, titrated carefully to avoid post-exertional malaise (60). However, other modifiable factors associated with brain age include sleep disruption, anxiety, and depression (59). Interventions that address these symptoms, such as cognitive behavioral therapy may protect against accelerated brain aging (61, 62). Future research is necessary to elucidate the protective mechanisms of healthy lifestyle habits on brain health and aging among patients with COVID-19.

COVID-19 mortality and morbidity are higher in men compared to women (63). Some studies have observed sex differences in COGVID (64) while others have not (65, 66). We found that males scored more poorly than females on a measure of attention and processing speed during a prior case series study involving a different sample at approximately 4 months post-infection (67). In the present cohort, at approximately 10 months post-infection, we found no sex differences in cognitive outcomes (18) but males had a 99% greater risk of accelerated BAG. The mechanisms of this sex difference remain unclear but may involve ACE2 pathways (68). For example, Swärd et al. showed that peripheral ACE2 expression is higher in males and increases more with age compared to females, at least in early adulthood (69). It is unknown if there is greater cerebral ACE2 upregulation in males, but it is plausible given the peripheral difference, and would potentially support ACE2 targeted therapies for this population.

We also found that there was a statistically significant decrease in risk of accelerated BAG associated with longer time since COVID-19 diagnosis. Given that this was only 1.4%, it may not be clinically meaningful. Alternatively, this finding may point to one or more subgroups of patients whose brain health significantly improves over time. If so, BAG may be useful in larger samples for biotyping (biologically stratifying) patients with different trajectories of brain health after COVID-19. Thus, BAG biotypes could improve precision medicine by identifying subgroups of patients at highest risk for COGVID (70). Evidence shows that a significant number of patients with post-COVID symptoms have considerable symptom resolution within a year after infection (71–73). Thus, longitudinal studies of COVID-19 survivors are needed to determine if BAG also improves over time and if so, in which patients.

Strengths of our study include the focus on mild to moderate COVID-19, whereas prior neuroimaging research has focused on post-hospitalization patients, the emphasis on younger survivors who may be more vulnerable to long-term cognitive effects, the novel examination of brain age in this population, the use of a well-established algorithm for estimating brain age, the inclusion of a control group, and the use of both objective and subjective cognitive assessments. Weaknesses include small sample, cross-sectional design which precludes insight regarding how participants functioned cognitively prior to COVID-19 and what individual cognitive trajectories may exist, self-reported nature of COVID diagnosis/symptoms, and the lack of data regarding lifestyle factors associated with brain aging (e.g., diet, smoking status) and other post-acute symptoms such as post-exertional malaise, ageusia, chronic cough, etc. that could be used to determine if any participants had other characteristics of Long COVID. There are several alternative algorithms available for estimating brain age that may yield different results, although we chose the one with the highest reliability based on the existing literature. Additionally, the brainageR algorithm utilizes gray, white, and CSF volumes whereas other algorithms use only gray matter. However, BAGs derived from different imaging modalities may represent different endophenotypes (23) and thus multimodal BAGs may be informative within larger, future COVID-19 studies.

In summary, our findings support our hypothesis that brain age is accelerated in survivors of COVID-19 infection compared to non-infected controls. This study represents the first examination of this innovative biomarker in post-COVID-19 neurocognitive dysfunction. Whereas samples from previous works have primarily included older adults, our study focused on a younger group of individuals to examine their specific cognitive vulnerabilities and associated neural mechanisms. Our cohort, observed on average after a longer duration post-COVID-19 infection than those in previous studies, offers distinct insights into the protracted effects of COVID-19 on brain health. Additionally, our findings contribute significant new information regarding the disparity in outcomes for men post-COVID-19.

## Data Availability

The datasets generated and analyzed during the current study are not publicly available due the fact that they constitute an excerpt of research in progress but will be available on the EBRAINS platform (https://www.ebrains.eu) under the corresponding author name at study completion. Interested parties may contact the corresponding author for further information.

## Acknowledgments

The authors would like to thank the faculty and staff of the Biomedical Imaging Center at The University of Texas at Austin.

## Author contributions

This study was developed by the principal investigator (SRK). Data acquisition, preparation and analysis was conducted by OFR, ADS and SRK. The manuscript was prepared and edited by OFR and SRK and reviewed and edited by all other authors. All authors approved the manuscript for submission. OFR, ADS and SRK had access to the raw data.

## Data Sharing

The datasets generated and analyzed during the current study are not publicly available due the fact that they constitute an excerpt of research in progress but will be available on the EBRAINS platform (https://www.ebrains.eu) under the corresponding author’s name at study completion. Interested parties may contact the corresponding author for further information.

